# A world apart: levels and factors of excess mortality due to COVID-19 in care homes. The case of Wallonia - Belgium

**DOI:** 10.1101/2020.08.29.20183210

**Authors:** Olivier J Hardy, Dominique Dubourg, Mélanie Bourguignon, Simon Dellicour, Thierry Eggerickx, Marius Gilbert, Jean-Paul Sanderson, Aline Scohy, Eline Vandael, Jean-Michel Decroly

## Abstract

COVID-19 became pandemic in 2020 and causes higher mortality in males (M) than females (F) and among older people. In some countries, like Belgium, more than half of COVID-19 confirmed or suspected deaths occurring in spring 2020 concerned residents of care homes. The high incidence in this population is certainly linked to its peculiar age structure but could also result from its poorer general health condition and/or from a higher contamination through the staff of care homes, while protection equipment and testing capacity were initially limited. To address these issues, we used data from Wallonia (Belgium) to characterize the distribution of death rates among care home institutions, to compare the dynamics of deaths in and outside care homes, and to analyse how age and sex affected COVID-19 death rates inside and outside care homes. We also used annual death rates as a proxy for the health condition of each population. We found that: (1) COVID-19 death rate per institution varied widely from 0‰ to 340‰ (mean 43‰) and increased both with the size of the institution (number of beds) and with the importance of medical care provided. (2) 65% of COVID-19 deaths in Wallonia concerned residents of care homes where the outbreak started after but at a faster pace than the outbreak seen in the external population. (3) The impact of age on both annual and COVID-19 mortality closely follows exponential laws (i.e. Gompertz law) but mortality was much higher for the population living in care homes where the age effect was lower (mortality rate doubling every 20 years of age increment in care homes, 6 years outside them). (4) Both within and outside care homes, the ratio of M/F death rates was 1.6 for annual mortality but reached 2.0 for COVID-19 mortality, a ratio consistent among both confirmed and suspected COVID-19 deaths. (5) When reported to the annual death rate per sex and age, the COVID-19 relative mortality was little affected by age and reached 24% (M) and 18% (F) of their respective annual rate in nursing homes, while these percentages reduced to 10% (M) and 9% (F) in homes for elderly people (with less medical assistance), and to 5% (M) and 4% (F) outside of care homes. In conclusion, a c. 130x higher COVID-19 mortality rate found in care homes compared to the outside population can be attributed to the near multiplicative combination of: (1) a 11x higher mortality due to the old age of its residents, (2) a 3.8x higher mortality due to the low average health condition of its residents, and (3) probably a 3.5x higher infection rate (1.6x in homes for elderly people) due to the transmission by its staff, a problem more acute in large institutions. Our results highlight that nursing home residents should be treated as a very specific population, both for epidemiological studies and to take preventive measures, due to their extreme vulnerability to COVID-19.

## Introduction

Since its appearance in China by the end of 2019, the COVID-19 pandemic is known to cause higher mortality in males than females and among older people (Wenham et al. 2020). In Europe and North America, COVID-19 has had a dramatic impact on people living in care homes (Comas-Herrera et al. 2020, ECDC 2020, Fisman et al. 2020, Ladhani et al. 2020, Petretto & Pili 2020), although one of the first policy applied to contain the outbreak was to ban family visits in care homes, usually at least one week before imposing containment restrictions to the whole population (Comas-Herrera, Ashcroft and Lorenz-Dant 2020, Verbeek et al. 2020). The high incidence of COVID-19 in care homes is likely related to the everyday contacts with the nursing personal, facilitating contagion (Arons et al. 2020, Ladhani et al. 2020), especially as many countries failed to provide sufficient personal protective equipment (PPE) for care homes at the beginning of the pandemic, usually concentrating the protections in limited supply (e.g. chirurgical masks and gloves) to hospitals (Logar 2020 for Italy, Rada 2020 for Spain, Quigley et al. 2020 for the United States, Szczerbińska 2020 for an international comparison). However, to assess if mortality rate due to COVID-19 was higher in care homes, it is important to factor out the age and sex effects, care homes being mostly populated by old people and also over-represented by women (Einiö et al. 2012). It is also important to take into account the generally lower health status of the population living in care homes (Falconer and O’Neill 2007), since people often enter care homes due to a deteriorating health status (Herm, Poulain and Anson, 2014) and that care home residents tend to have a lower social status than the rest of the population (Laferrère et al. 2013). Finally, distinguishing different categories of care homes according to their size or the types of health assistance they provide to their residents could be instructive to compare how COVID-19 affected populations differing by their general health conditions and/or by the risk of contamination through nursing services.

As PCR testing capacities were also strongly limited at the beginning of the outbreak, there was often a lack of test to assess whether deaths occurring within care homes resulted from COVID-19. In some countries like Belgium, the reporting of COVID-19 deaths by national authorities has been inclusive, including people suspected to have died from COVID-19 due to characteristic symptoms despite the absence of PCR testing (Comas-Herrera et al., 2020). The excess of deaths in spring 2020 compared to previous years according to national register data gave clear support to this inclusive approach (Sciensano 2020c; *Bustos Sierra et al., submitted)*. From 8 March to 9 May 2020, during the most intense phase of the epidemic in terms of mortality, 8,735 COVID-19 deaths (confirmed and suspected) were recorded in Belgium, while over the same period an excess mortality of 8,280 deaths was recorded in the National Register (a difference of 455 deaths, barely 5% less) compared to the average mortality of 2015-19. However, it has been suggested that social isolation and the stress caused by anxiety could strongly affect elderly people during lockdown (Armitage & Nellums, 2020) and might also have been responsible for a substantial portion of excess mortality in nursing homes (Trabucchi and De Leo 2020). Although it is extremely hard to identify retrospectively the cause of death, we can expect that if COVID-19 mortality rate is affected by age and sex following a particular pattern, COVID-19 suspected deaths should display the same pattern as COVID-19 confirmed deaths whenever the diagnose was correct.

Our objective is to characterize in Wallonia (southern Belgium) mortality resulting from COVID-19 in different populations: residents of care homes for elderly people and the rest of the population, while accounting for age and sex effects as well as the general health status of each population. First, we show the overall contrasts in mortality by COVID-19 between populations living in and out of care homes, as well as the variations in mortality between care home institutions according to the importance of medical assistance (distinguishing nursing homes from residential homes) and their size. We also compare the temporal dynamics of mortality in and out of care homes because we hypothesize (i) that the outbreak in care homes started after the one occurring outside care homes if infections originated from the nursing personal, and (ii) that the potentially high contagion within care homes might have caused a more rapid spread of the virus. Second, we seek to explain the mortality differential within and outside care homes through two factors: differences in structure by age and sex (structure effect) and differences in COVID-19 age and sex-specific death (behavioral effect). After highlighting these two effects, we attempt to explain the behavioral effect, which results from (i) the unequal incidence of the disease in the populations being compared and (ii) the greater or lesser vulnerability of individuals once they have contracted the SARS-CoV2. The latter, which corresponds to more or less deteriorated health status, can be approximated by the usual level of mortality, outside of the COVID-19 health crisis, in the populations studied. In the absence of systematic testing, it is not possible to measure the unequal incidence of the disease in the populations compared. However, we show that it is possible to estimate it by comparing mortality caused by COVID-19 and annual mortality because the two are tightly correlated in each population. Through these different analyses we provide a better understanding of the key factors at the origin of the high incidence of COVID-19 on the mortality in care homes.

## Materials and Methods

### Area of interest and institutional context related to care homes

Due to data availability of both COVID-19 and annual deaths, we focus on the French-speaking part of Wallonia in southern Belgium (i.e. the Walloon Region without the districts attached to the German-speaking community of eastern Wallonia). In this area, comprising c. 3.6 million inhabitants, all residential facilities for elderly people (RFEP)^1^ are supervised by the regional administration AViQ (“Agence pour une Vie de Qualité”, https://www.aviq.be) who collected statistics regarding COVID-19 deaths, and performed recurrent systematic surveys on yearly mortality, the last one occurring in 2017. For some analyses, among RFEP we distinguish homes for elderly people (HEP), which host essentially old people who require assistance for daily meals, housekeeping and/or daily toilet but do not require substantial health care, and nursing homes (NH), which host essentially old people with the same needs as HEP residents but also people requiring additional health care due to more or less severe pathologies. According to the standards established by the *Code réglementaire wallon de l’Action sociale et de la Santé*, the number of nursing staff for 30 residents is 4.5 in HEP against 12.1 in NH. The latter must also have at least five nurses, one coordinating doctor and 0.1 full-time equivalent specialist in palliative care. NH also have written procedures for hand hygiene and the isolation of residents with an infection that carries a risk of contamination. On average, approximately half of the bed capacities of NH is devoted to host residents requiring health care services. The Walloon population not living in RFEP will be collectively referred to as the unassisted population (UA), because it is essentially composed of people living autonomously, although a small proportion lives in very particular contexts (e.g. prisons).

### COVID-19 mortality data

Our analysis of mortality by COVID-19 in and out of RFEP covers the period between March 13 and June 30, 2020. The data we use came from two sources. First, AViQ collected death data attributed to COVID-19 as reported by all 573 RFEP (446 NH and 127 HEP) existing in 2020 (data including death date, age and sex, which concerned only long-term residents). These deaths occurred essentially in hospitals (28% of cases), in which case a PCR and/or scanner test confirmed the COVID-19 diagnostic, or within NH or HEP institutions (72% of cases) where tests were available for only 27% of cases so that the majority of these cases are suspected to result from COVID-19, following the diagnosis of the attending physician and given the presence of characteristic symptoms (upper or lower respiratory tract infection, fever or chills, cough, shortness of breath or difficulty breathing, headache, new loss of taste or smell, etc; Dequeker et al. 2020). Second, Sciensano, the Federal Institution responsible for Health monitoring in Belgium (https://www.sciensano.be), collected COVID-19 related deaths data reported by hospitals and other settings, such as the agencies surveying car homes (including AViQ). We extracted from their dataset all COVID-19 related deaths (suspected and confirmed) reported in Wallonia and applied a correction to account that our focal area does not include 2.1% of the Walloon population (German-speaking districts). It should be noted that the Sciensano dataset considers the place of death, rather than the place of residence, to distribute deaths among Belgian regions. However, we estimated that the deaths of Walloons that occurred outside (e.g. in hospitals from the Brussels-Capital Region) were nearly compensated by non-Walloons who died in Wallonia (they represented c. 1% of the deaths according to partial data).

To assess the heterogeneity of COVID-19 mortality among institutions, the crude COVID-19 death rate per institution was computed by dividing the number of reported deaths by the total bed capacity of the institution. This ratio underestimates the actual mortality rate but should remain realistic given that >90% of the bed capacity was usually occupied at the beginning of the outbreak. By institution, we here considered a physical site that received a particular approval number in the AViQ database, although some sites are managed by the same administrative entity.

To characterize the temporal dynamics of COVID-19 deaths from 13 March until 30 June 2020, we computed the cumulated number of deaths for RFEP residents and for the UA population (by subtracting the AViQ data from the Sciensano data for Wallonia). To assess the delay and pace of the outbreaks that occurred in each population, we also computed the median date at which people from each population died (i.e. when 50% of all deaths occurred) as well as the dates corresponding to the 0.05 and 0.95 quantiles.

To assess the age and sex effects, COVID-19 death rates were computed per sex and 5-years age classes, separately for the RFEP and the UA populations, as the ratio of the number of COVID-19 deaths over the corresponding age and sex-specific population sizes estimated on January 1, 2020. For RFEP, these population sizes were derived from the population survey established on January 1, 2018, by AViQ (see below), after applying a multiplication factor reflecting the change in total bed capacities between January 1, 2020, and January 1, 2018 (factor of 42103/40852 = 1.031 for NH and 6249/6865 = 0.910 for HEP). For the UA population, we used the StatBel data (https://statbel.fgov.be), based on the National Register, which provided the population size disaggregated by age and sex for our focal area on January 1, 2020, and subtracted the RFEP population.

### Annual mortality data in 2017

We compared the overall health status of each population using their respective age and sex-specific annual death rates before the COVID-19 epidemic. AViQ conducted a survey in 2017 asking each of the 587 RFEP (440 NH and 147 HEP) existing that year to report deaths (including birth and death dates) throughout the year, and to report their population on January 1, 2018. Here, we considered only deaths and population of long-term residents, excluding people that registered for revalidation short-stays (typically less than a month) in some of these facilities because such services were closed during the 2020 sanitary confinement period. Overall, in terms of bed capacities, 93.3% of NH and 84.8% of HEP provided their population statistics, and respectively 91.3% and 83.2% reported their 2017 deaths. Sex and age data were available for at least 98% of the people in each case. To correct raw numbers, they were divided by the proportion of responding institutions (in bed equivalents) and by the proportion of complete sex and age data, so that incomplete data were reported across all age by sex categories.

We extracted from StatBel population statistics the population on January 1, 2018, as well as the number of deaths during 2017, disaggregated by age and sex, for our focal area. From this overall population and death numbers, we subtracted the corresponding estimated numbers for RFEP (based on the AViQ 2017 survey) to estimate the population and associated deaths of people living outside of care homes, i.e. the UA population.

Annual death rates were then computed for 5-years age categories by sex, separately for each population (UA and RFEP in which we also distinguished NH and HEP), by dividing the number of deaths occurring in 2017 by the population size on January 1, 2018.

### Data analyses

The distribution of COVID-19 death rate per institution was established separately for NH and HEP, and within each type of institution separately for the smallest and largest institutions in terms of bed capacity. The criterion used to distinguish small and large institutions is where approximately half of the residents of a particular institution type (NH or HEP) live. Mann-Whitney U tests were used to assess whether the medians of these distributions differ significantly.

We compared the age pyramids of the RFEP and UA populations using population data on January 1, 2020. To assess to which extent the age pyramid of RFEP residents affect its COVID-19 death rate, we predicted the crude COVID-19 death rate of this population using UA age and sex-specific COVID-19 death rates weighted by the age and sex-specific RFEP population sizes, and compared it to the crude COVID-19 death rate observed in the UA population.

For each sex and population, COVID-19 age-specific death rates per 5-years classes (m) were computed. Because the size of the HEP population is relatively small, age-specific COVID-19 death rates often lack sufficient precision, so that we generally report them for RFEP (i.e. NH + HEP) only, and highlight differences between NH and HEP on overall mortality rates because the two populations have very similar age pyramids.

COVID-19 age and sex-specific death rates were represented on a log scale against the mid-age (a) of each class, and adjusted on an exponential function (i.e. Gompertz’ law) restricted to age classes between 65 and 99 years (to avoid classes with too small population sizes): *m* = *c.e^b.a^*, where *b* and *c* are the adjusted coefficients. We estimated the number of years leading to a doubling of *m* as ln(2)/b. We used Fisher distribution to compute 95% confidence intervals of mortality rates following Ars *et al*. (1988) as the interval between 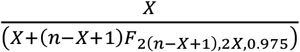 and 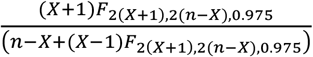 where *X* and *n* represent the age and sex-specific death number and population size, respectively, and *F_d1,d2,α_* represent Fisher’s distribution at level *α* with *d*1 and *d*2 degrees of freedom. The same procedure was applied on annual mortality rates.

To assess to which extent the health condition of RFEP residents is lower than that of the UA population at the same age and sex, we compared the crude annual death rate observed in the RFEP population with the one predicted using UA age and sex-specific annual death rates weighted by age and sex RFEP population sizes. Finally, to attempt to factor out the effect of the general health condition of a particular population on COVID-19 mortality, we divided age and sex-specific COVID-19 death rates by the respective annual death rates. We then compared these ratios among populations to tentatively interpret them in terms of relative COVID-19 infection rates.

### Interpretation of death rates

Computing classical demographic rates on mortality in RFEP is complicated due to the high turnover of people and the availability of death data for only a single year. However, with an increase of overall bed capacities of only 0.6% between 2017 and 2018 and the fact that c. 98.3% of beds are occupied, we can consider that the RFEP population is stable during a single year. Therefore, while the annual mortality rates computed overestimate the probability that a RFEP resident dies within a year, they can be interpreted as 365 times the probability that a RFEP resident dies within a day. They are thus proportional to the instantaneous rate of mortality (the so-called force of mortality) and as such represent adequate measures of the average health condition of a particular age and sex category.

The situation is different for COVID-19 mortality rates because once the epidemics started in March 2020, RFEP institutions usually did not reintegrate new residents to replace their free beds until mid-June. We also ignore how many residents left their institution and were host in their family. Hence, COVID-19 death rates in RFEP must be somewhat underestimated.

## Results

Between March 13 and June 30, 2020, COVID-19 caused a total of 2,126 deaths in RFEP (2010 in NH, 116 in HEP) and c. 1,186 deaths in the UA population of the French-speaking part of Wallonia (Table 1). Hence, 65% of the suspected and confirmed deaths of COVID-19 occurred in RFEP, even though the population living in these institutions constitutes barely 1.3% of the Walloon population. Consequently, the crude COVID-19 death rates are extremely contrasted. They reach 44.2‰ in RFEP against 0.33‰ in UA, i.e. a ratio of 1 to 134. In Wallonia, as in other countries where care homes have been heavily impacted by COVID-19, two distinct epidemics have therefore developed, one in care homes and the other outside them (see Humblet 2020 for Belgium; Logar 2020 for Italy; Rada 2020 for Spain).

**Table 1.** Descriptive statistics about COVID-19 outbreak in Wallonia (excluding the German-speaking districts). Population (January 1, 2020) (A), COVID-19 deaths from march 13 to June 30 2020 (B) and crude COVID-19 death rates (per 1,000 people) (C), in residential facilities for elderly people (RFEP), including nursing homes (NH) and homes for elderly people (HEP), and in the unassisted population (UA). Statistics given for the whole population and the population aged 65 and more. Data sources: StatBel, AViQ.

| A : Population (January 1, 2020, in thousands) |  |  |  |  |  |  |  |  |
| --- | --- | --- | --- | --- | --- | --- | --- | --- |
|  | Total |  |  |  | 65+ |  |  |  |
|  | Both sexes |  | Females |  | Both sexes |  | Females |  |
|  | x 1,000 persons | % | x 1,000 persons | % both sexes population | x 1,000 persons | % total population | x 1,000 persons | % both sexes population |
| RFEP | 48 | 1.3 | 36 | 74.6 | 46 | 95.1 | 35 | 76.2 |
| NH | 42 | 1.2 | 31 | 74.5 | 40 | 95.2 | 30 | 76.1 |
| HEP | 6 | 0.2 | 5 | 74.7 | 6 | 94.2 | 5 | 76.4 |
| UA | 3,519 | 98.7 | 1,788 | 50.8 | 626 | 17.8 | 349 | 55.7 |
| Wallonia | 3,567 | 100.0 | 1,824 | 51.1 | 672 | 18.8 | 384 | 57.1 |

| B : COVID-19 deaths (march 13 to june 30, 2020) |  |  |  |  |  |  |  |  |
| --- | --- | --- | --- | --- | --- | --- | --- | --- |
|  | Total |  |  |  | 65+ |  |  |  |
|  | Both sexes |  | Females |  | Both sexes |  | Females |  |
|  | persons | % | persons | % of both sexes deaths | persons | % of total deaths | persons | % of both sexes deaths |
| RFEP | 2,126 | 64.6 | 1,354 | 63.7 | 2,090 | 98.3 | 1,339 | 64.1 |
| NH | 2,010 | 61.1 | 1,276 | 63.5 | 1,975 | 98.3 | 1,261 | 63.8 |
| HEP | 116 | 3.5 | 78 | 67.2 | 115 | 99.1 | 78 | 67.8 |
| UA | 1,163 | 35.4 | 452 | 38.9 | 974 | 83.7 | 392 | 40.2 |
| Wallonia | 3,289 | 100.0 | 1,806 | 54.9 | 3,064 | 93.2 | 1,731 | 56.5 |

| C : Crude COVID-19 death rates (per 1,000 people) |  |  |  |  |  |  |
| --- | --- | --- | --- | --- | --- | --- |
|  | Total |  |  | 65+ |  |  |
|  | Both sexes | Females | Males | Both sexes | Females | Males |
| RFEP | 44.2 | 37.8 | 63.1 | 45.7 | 38.5 | 68.9 |
| NH | 48.1 | 40.9 | 69.0 | 49.6 | 41.6 | 75.1 |
| HEP | 18.5 | 16.7 | 24.0 | 19.5 | 17.3 | 26.6 |
| UA | 0.3 | 0.3 | 0.4 | 1.6 | 1.1 | 2.1 |
| Wallonia | 0.9 | 1.0 | 0.9 | 4.6 | 4.5 | 4.6 |

### Distribution of COVID-19 death rate per institution

Large variations in mortality by COVID-19 are also observed between RFEP institutions, depending on their category and size. With 2,100 deaths out of 2,126 (95%), NH recorded a crude COVID-19 death rate (48‰) 2.6 times higher than HEP (19‰) (Table 1). COVID-19 death rate per institution varied widely from 0‰ to 340‰ and was affected by both the type of institution and their size (Fig. 1). About half of the residents of HEP lived in small-sized institutions ranging from 14 to 52 beds (mean = 37.8, N = 84) where 16‰ died from COVID-19, a lower rate than the 21‰ who died in medium-sized HEP institutions ranging from 54 to 148 beds (mean = 74.3, N = 43). Similarly, about half of the residents of NH lived in medium-sized institutions ranging from 29 to 104 beds (mean = 76.2, N = 284) where 40‰ died from COVID-19, a lower rate than the 53‰ who died in large-sized NH institutions ranging from 105 to 298 beds (mean = 134.1, N = 163).

**Figure 1.**
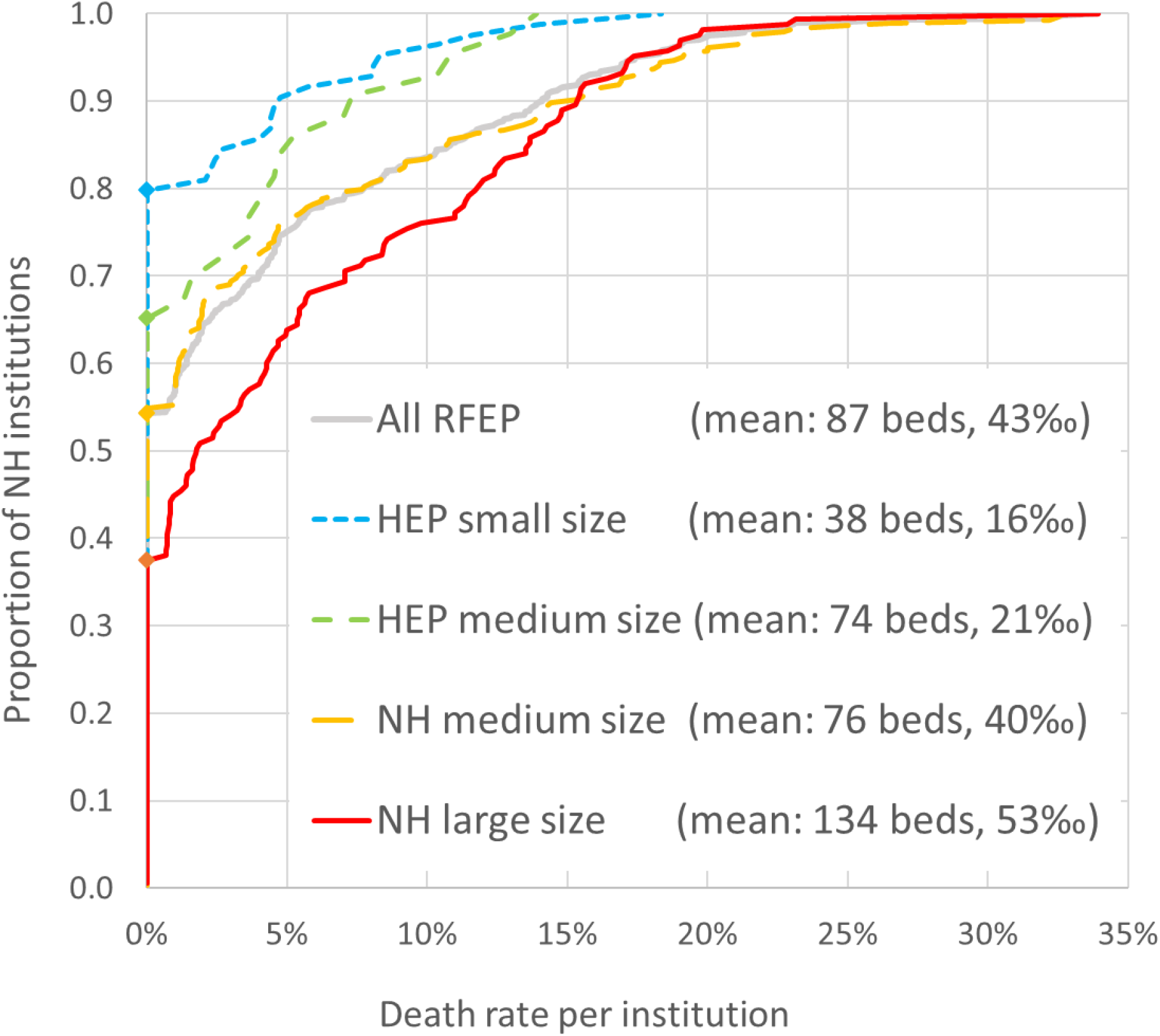
Cumulated distributions of COVID-19 death rate per RFEP institution (grey line) distinguishing nursing homes (NH) and homes for elderly people (HEP) and two size classes per RFEP category based on bed capacities (mean bed capacity indicated in the graph legend). The area above each curve is proportional to the overall death rate (indicated in the graph legend). The symbols along the vertical axis highlight the proportions of institutions where no death was reported. Data source: AViQ.

Remarkably, a significant proportion of RFEP (55%) recorded no death, showing that the epidemic has affected care homes in a very variable manner. The proportion of institutions without COVID-19 death followed the trend shown above for death rates: 81% and 67% for small and medium-size HEP institutions, compared to 55% and 38% for medium-size and large NH institutions, respectively (Fig. 1). Finally, while the crude COVID-19 death rate rarely exceeded 150‰ in HEP institutions, it exceeded this threshold in about 10% of NH institutions, regardless of size (Fig. 1). Mann-Whitney U tests on crude COVID-19 death rate per institution showed that globally NH and HEP differ significantly (z-score = 4.67, p-value < 0.00001) as well as medium NH versus large NH (z-score = -3.07, p-value = 0.001), while difference was only marginally significant between small HEP and medium HEP (z-score = -1.30, p-value = 0.097) and between medium HEP and medium NH (z-score = 1.20, p-value = 0.11).

### COVID-19 death temporal dynamics in and out of RFEP

While mortality levels by COVID-19 are significantly higher in RFEP than in UA, the temporal dynamics of mortality also differed in the two populations. Until April 5 the majority of deaths had occurred in the UA population while RFEP residents constituted the majority of cumulated deaths since April 6 (Fig. 2). The slight delay in the mortality dynamics of RFEP residents can be quantified by the date at which 50% of the deaths reported in the observed period occurred: 3.5 days later in the RFEP population (April 13) than in the UA population (April 10). The wave of deaths was also more concentrated in RFEP, with 52 days between the dates for which 5% and 95% of the total deaths were reached (i.e. between 28/03 and 19/05), than in the UA population (64 days between 23/03 and 27/05). Hence, according to mortality data, the outbreak that occurred within the RFEP was delayed but then spread faster than the outbreak seen in the UA population.

**Figure 2.**
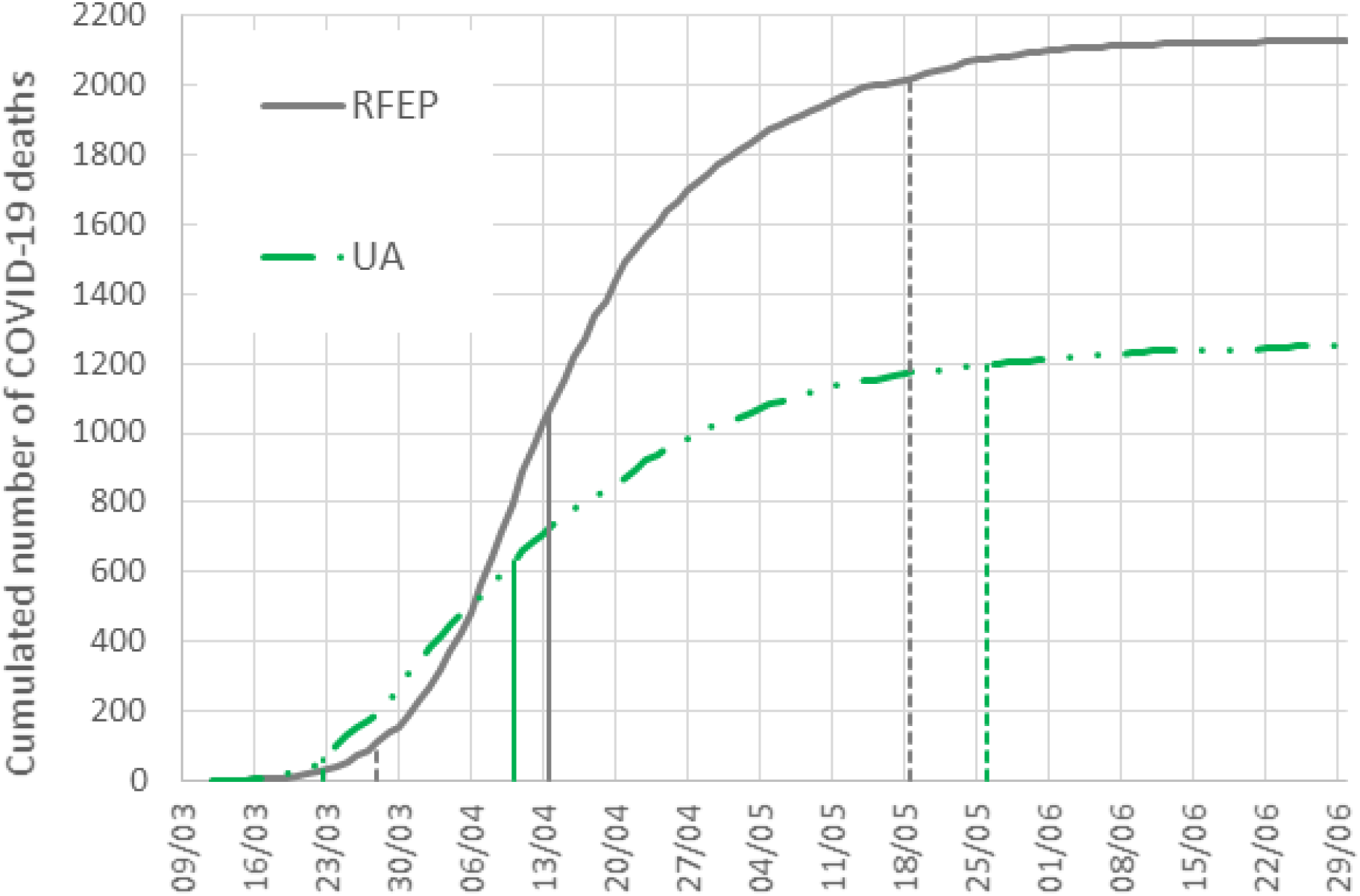
Cumulated number of deaths attributed to COVID-19 in Wallonia between March and June 2020, separately for persons living in residential facilities for elderly people (RFEP, grey plain line) and the unassisted population (UA, green dotted-stippled line). The dates at which 50% of the total deaths recorded in each population are indicated by vertical bars, while the 0.05 and 0.95 quantiles are indicated by vertical stippled lines. Data sources: Sciensano, AViQ.

### Age and sex structure effect

The RFEP population, which makes 1.35% of the French-speaking part of Wallonia (3,567,294 inhabitants), has a peculiar age structure, with people aged 65 and over constituting 95% of the total population, compared to 18% in UA (Fig. 3). Moreover, among the people aged 65 and over, the oldest (85 and over) are over-represented in care homes, accounting for 60% compared to only 11% in UA. As a result, the mean age reaches 84.2 years (SD = 9.6) in the RFEP and only 41.0 years (SD = 23.2) outside them. The gender structure is also very particular in the RFEP, characterized by a highly biased sex ratio, with 74.6% of females (51.1% in the overall population), a percentage increasing from 50.9% among the 60-64 years old to 91.2% in the ≥100 years old. Within the RFEP, the age pyramids of residents of NH and HEP are very similar, the main difference being a slightly higher proportion of males 60-74 years old in HEP (8.0%) compared to NH (6.3%), compensated by a lower proportion of males 80-94 years old in HEP (12.2%) compared to NH (13.8%).

**Figure 3.**
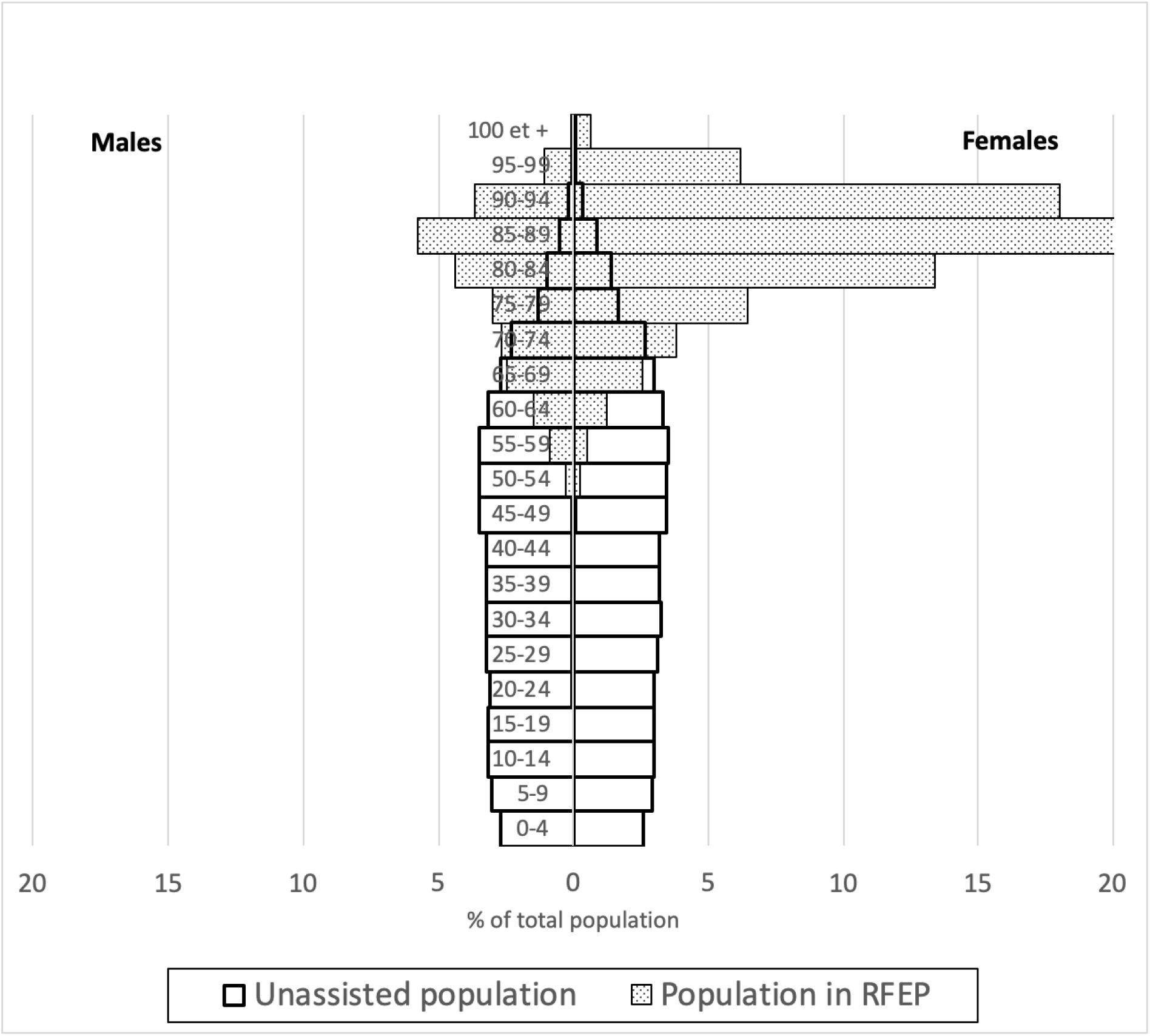
Age pyramids (in %) of the populations of the French part of Wallonia on January 1, 2020. People living in residential facility for elderly people (RFEP, in grey) and unassisted population (UA, white). The 65+ people made 95.1% of the RFEP population and 17.8% of the UA population. Data sources: StatBel, AViQ.

Given the strong excess of females among RFEP residents, especially among the oldest and more vulnerable age classes, females constitute a majority of the COVID-19 deaths in RFEP. By contrast, in the UA population composed of 51% females, 39% of COVID-19 deaths were females. Overall, in Wallonia, females constituted 55% of COVID-19 deaths despite their lower vulnerability (see below) because a much higher proportion of females ≥65 years old live in RFEP (9.2%) than males ≥65 years old (3.9%).

The age and sex-specific COVID-19 death rates observed in the UA population (see below) weighed by the RFEP population sizes predicted a crude death rate of 3.55‰ in RFEP, which is 10.7 times higher than the observed crude COVID-19 death rate in the UA population (0.33‰). Hence, the difference of age pyramids between the RFEP and UA populations, i.e. the structure effect, already predicts an 11-fold difference in their overall COVID-19 death rates.

### COVID-19 death rates by sex and age groups – behaviour effect

In RFEP and in UA, age and sex-specific death rates due to COVID-19 (confirmed and suspected) closely followed Gompert’s law (i.e. exponential increase with age), but with a lower slope in the former (mortality doubling every 21.5 years in M and 19.9 years in F) than in the latter (mortality doubling every 6.3 years in M and 6.4 years in F) (Fig. 4A). COVID-19 death rates were higher in males than in females of the same age class, by a factor of c. 2.0 in RFEP and 2.1 in UA among the 65-99 years old people. As already shown with the distribution of death rates per institution (Fig. 1), there was also a sharp contrast between NH and HEP populations: across all ages, COVID-19 death rates in NH (6.9% in M, 4.1% in F) were approximately 2.9 (M) to 2.5 (F) times higher than in HEP (2.4% in M, 1.7% in F).

**Figure 4.**
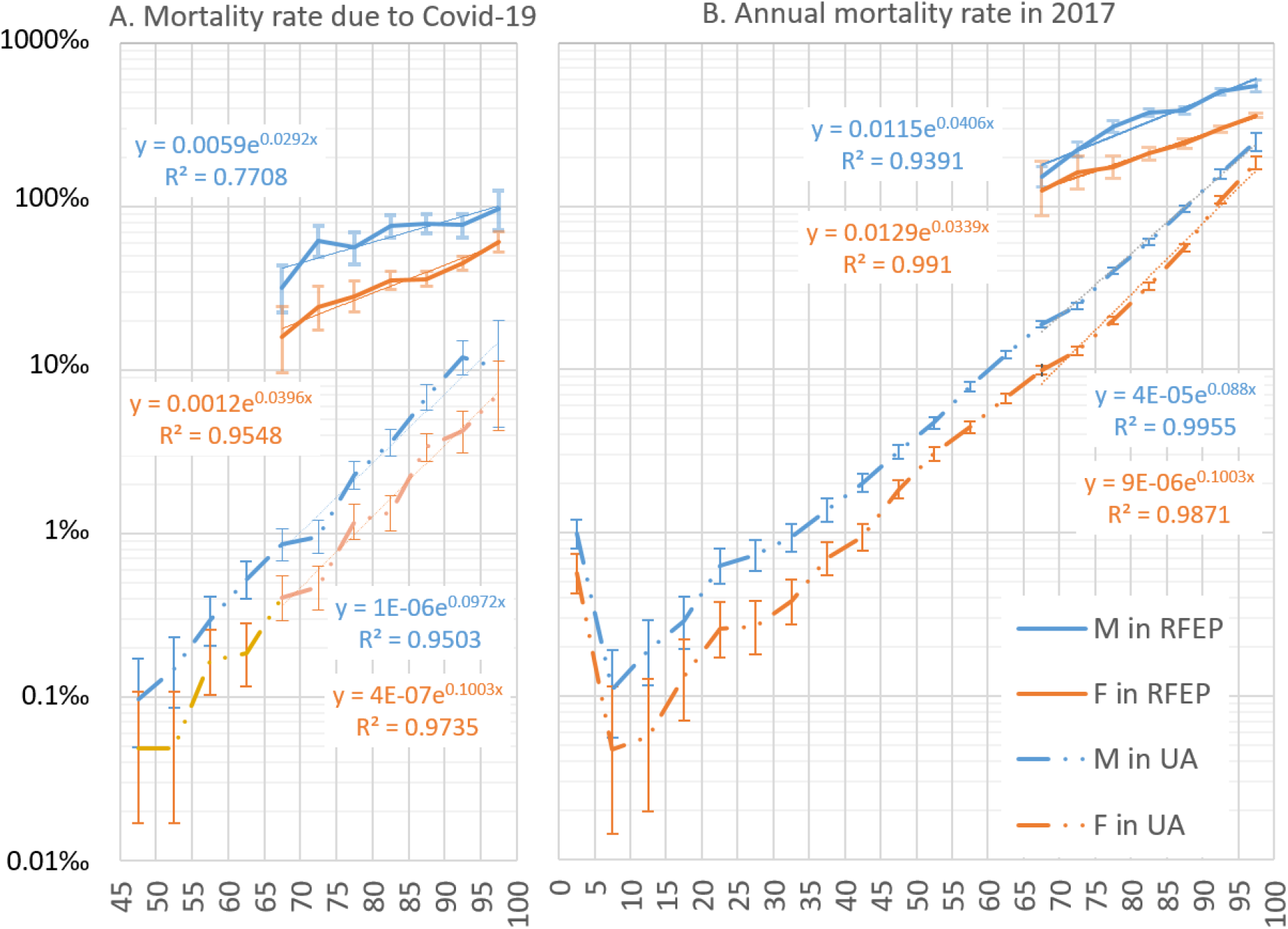
Age and sex-specific death rates due to COVID-19 during spring 2020 (**A**) and due to all causes during 2017 (**B**) in Wallonia, separately for residential facilities for elderly people (RFEP, plain lines) and the unassisted population (UA, stippled-dotted lines), and for males (blue) and females (red). Death rates are represented on a logarithmic scale. Equations and thin straight lines represent the best-fitting exponential curves adjusted to the 65-99 age classes. Data sources: Sciensano, AViQ, StatBel.

At all ages, COVID-19 death rates were significantly higher in RFEP than outside them (Fig. 4A), reflecting the behaviour effect. However, the ratio between RFEP over UA rates tends to decrease with age: while it exceeds 30 for people under 75, it reaches 10 for those aged 85-89 (the age group concentrating most COVID-19 deaths, both inside and outside RFEP), then 3 for people over 99 years. Thus, the mortality differential is most pronounced at the youngest ages. To measure the extent of the contrast in mortality by COVID-19 between care homes and the rest of the population, it should be noted that if the population had been subjected to the age and sex-specific COVID-19 death rates observed in RFEP, there would have been more than 38,500 deaths in UA (compared to 1,163).

**Figure 5.**
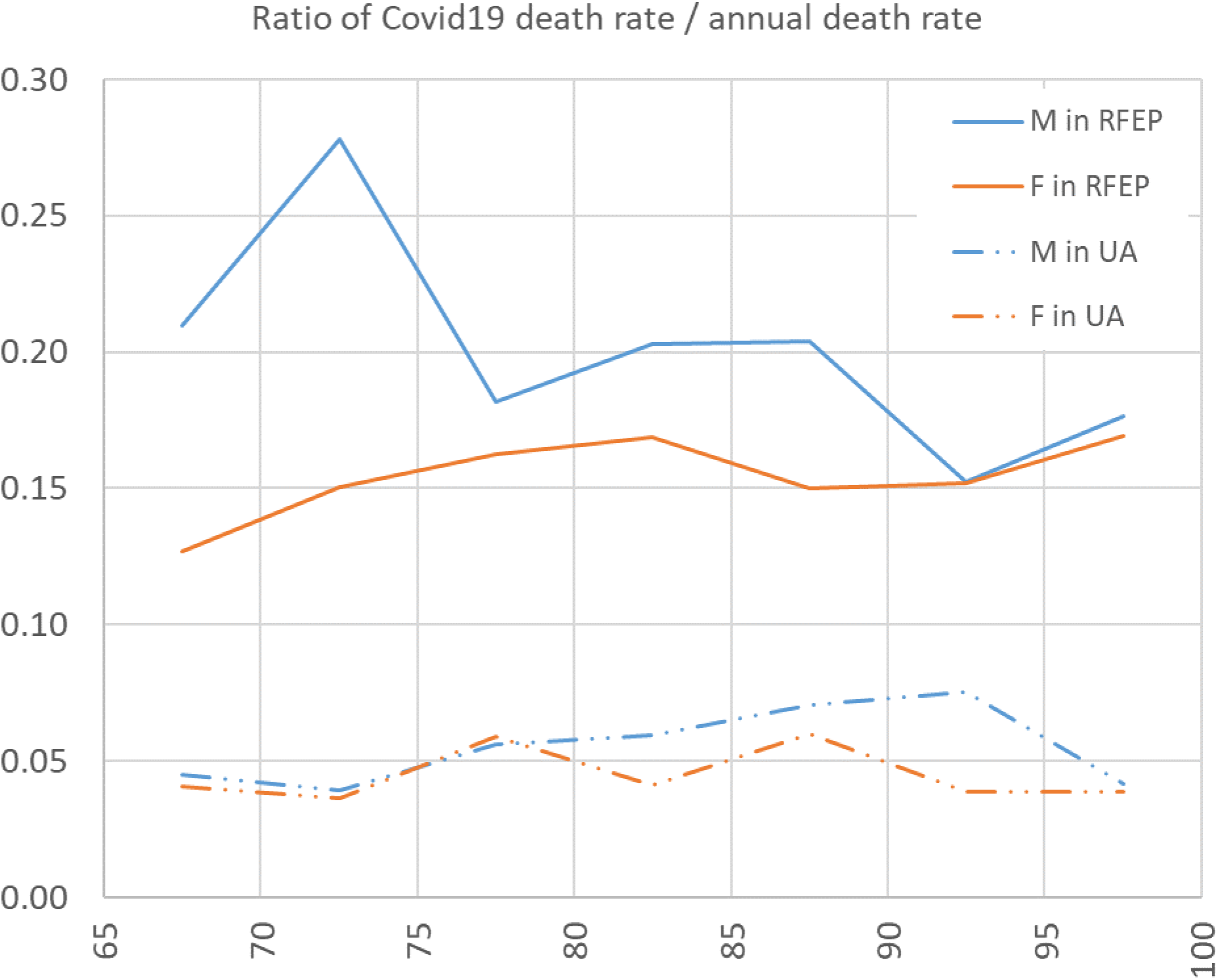
Ratio of COVID-19 over annual death rates in males (M, blue) and females (F, red) separately for residential facilities for elderly people (RFEP, plain lines) and the unassisted population (UA, dotted-stippled lines). Data sources: AViQ, Sciensano, StatBel.

One way to measure the contribution of differences in age and sex-specific death rates between RFEP and UA, i.e. the behaviour effect, is to compare the crude COVID-19 death rate in RFEP with the one predicted by applying the age and sex structure of RFEP to the age and sex-specific death rates of UA. As already indicated, the first rate reaches 44.2‰ and the second 3.55‰. The ratio between the two is therefore 12.5. In other words, the excess mortality observed in RFEP results from the combination of a structure effect (x 10.7) and a behaviour effect (x 12.5), their product giving the ratio between the crude COVID-19 death rates of the two populations (10.7 x 12.5 ≈ 134).

### Annual overall death rates by sex and age groups - a proxy of overall health status

When excluding infantile mortality (0-4 years class), age and sex-specific death rates based on 2017 mortality data also closely followed Gompertz’s laws, both in the RFEP and UA populations, but were overall 5 (in RFEP) to 20 (in UA) times higher than the COVID-19 death rates (Fig. 4B). Age-specific death rates were higher in males (M) than in females (F) of the same age class, by a factor of c.1.6 in RFEP and 1.7 in UA among the 65-99 years old people. However, mortality rates were much higher in RFEP than in UA for the same age class and they increased with age less rapidly in RFEP (mortality doubling every 15.1 years in M and 17.8 years in F) than in UA (mortality doubling every 7.7 years in M and 6.6 years in F). Therefore, while annual death rates were c. 10 times higher in RFEP than in UA for the 65-69 class, they remained only c. 2 times higher for the 95-99 class (Fig. 4B), indicating that the morbidity of care home residents tends to be stronger among the youngest residents when their age is taken into account.

The overall annual death rates over all age classes and sex reached 266.7‰ in RFEP against 7.0‰ in the UA populations, a 38-fold difference. Using the age and sex-specific death rates observed in UA weighed by the population sizes of the RFEP population predicts a crude death rate of 70.7‰ in the RFEP population if it was as healthy as the UA population under a same age pyramid. Hence, the lower health condition of the RFEP population can explain a 266.7/70.7 = 3.77-fold increase in mortality compared to the UA population. Across all age classes, annual death rates in NH (34.1% in M, 25.2% in F) were approximately 1.2 times higher than in HEP (27.6% in M, 20.1% in F), reflecting that NH institutions host a higher proportion of residents in poor health condition than HEP institutions.

### Comparison of COVID-19 and annual age and sex-specific death rates

While mortality levels by age and sex of COVID-19 are significantly lower than those of 2017, within each population (RFEP, AU) and sex the COVID-19 death rate is nearly proportional to the annual death rate. Indeed, in both cases, mortality is systematically higher in RFEP than outside, it increases with age, but with a lower slope in care homes and a strong excess of male mortality is observed.

However, there are two major differences between COVID-19 and annual mortality. First, the gap between mortality by COVID-19 and annual mortality is lower in care homes than in the rest of the population. Consequently, the contrast in death rates between the RFEP and UA populations after accounting for sex and age effects were even higher than for the annual mortality: COVID-19 death rates were c. 30 times higher in RFEP than in UA for the 65+ class, with a decrease of the ratio with age (c. 38 times higher for the 65-69 class to c. 9 times higher for the 95-99 class) (Fig 4A). Second, the excess mortality in males, already present in the annual overall mortality, is even higher in the case of COVID-19 mortality. In and out care homes, while the male-to-female mortality ratio for those aged 65+ reached c. 1.6 in 2017, it exceeded 2.1 for COVID-19 mortality.

### Relative impact of COVID-19 on the mortality of the different populations and their contamination

The fact that within each population (RFEP, UA) and sex the age specific COVID-19 death rate is near proportional to the annual death rate (Fig. 4a,b) suggests that the latter might be a good proxy of the average effect of comorbidity factors conditioning the risk of dying from COVID-19 when one is infected. Under this assumption, we can tentatively factor out the impact of comorbidity factors using the ratio of COVID-19 over annual death rates for each age class and sex (Fig. 5). Although these ratios are slightly higher for males than females in both RFEP and UA populations, and a little higher before age 75 than at later ages for men in RFEP, the main difference is between populations: while COVID-19 death rates are around 16% (F) to 20% (M) of the annual death rates in REFP, they reached only 5% (F) to 6% (M) of the annual death rates in UA. If we assume that this difference results from different levels of exposure to the SARS-CoV-2 virus causing COVID-19, REFP residents would have been c. 3.5 times more contaminated than the UA population of similar age. Moreover, average ratios over the 65-99 old people distinguishing NH from HEP populations indicate that the contamination was probably 1.6 (HEP) to 3.8 (NH) times higher than in the UA population, and thus that NH institutions would have been at least twice more contaminated than HEP institutions.

## Discussion

In Wallonia, care home residents constitute only 1.33% of the population but 65% of the COVID-19 deaths reported during spring 2020, so that the death rate was c. 130 times higher than for the population living outside of care homes. According to our analyses, this seemingly dramatic ratio results from the combination of three main factors. First, the peculiar age pyramid of the care homes population predicts a crude COVID-19 death rate 11 times higher than for the external population, because COVID-19 is much more deadly for older people (structure effect). Second, the low average health condition of care home residents increases their annual death rate by c. 3.8 times compared to the external population after accounting for age and sex effects, while COVID-19 mortality appears highly correlated to the presence of other comorbidity factors (behaviour effect resulting from lower health status). Finally, care home residents have probably been on average 3.5 times more exposed to the SARS-CoV-2 virus than the external population of similar age (1.6 times more in HEP, 3.8 times more in NH), and tended to be more exposed when living in larger institutions (behaviour effect resulting from higher SARS-CoV-2 prevalence). All these effects worked nearly multiplicatively to generate such a high death toll on care homes. These inferences are based on a number of assumptions that merit to be discussed.

The higher mortality of COVID-19 on older people has been reported since the beginning of the epidemics (Verity et al. 2020). Our analyses using 5-years age classes allow us to better characterize this relationship, showing that the death rate doubles approximately every 6 years of age increment for people living outside of care homes, and every 20 years for care home residents, a result similar to that observed by Guilmoto (2020) in a comparison of age and sex-specific death rates in Western Europe and the United States. This type of mortality-age relationship is known as Gompertz law (Gompertz 1825), which is sometimes interpreted as reflecting an increase in the vulnerability of individuals to extrinsic causes (Ricklefs & Scheuerlein 2002). Gompertz law often fits well mortality data for a wide range of causes (Riggs 1991 for ischemic heart disease in the US; Imaizumi 1996 for breast cancer in Japan), as well as total mortality rates at national level (all causes confounded, Horiuchi *et al*., 2003; Dolejs 2015), as confirmed in our analyses of the annual mortality rate (if we exclude infantile mortality). What is remarkable here is that the exponents of the Gompertz law adjusted to our data were very similar for COVID-19 mortality and overall annual mortality (Fig. 4A, B) while they differed strongly between populations living inside or outside care homes. This strong correlation between COVID-19 and annual mortality rates suggests that the risk of dying from COVID-19 largely depends on the person’s initial health condition, as supported by numerous clinical observations showing the strong impact of multiple comorbidity factors (Williamson *et al*. 2020).

The higher mortality of COVID-19 on males than females has also been reported since the beginning of the epidemics (Verity et al. 2020; Wenham *et al*. 2020; Williamson *et al*. 2020). In Wallonia, COVID-19 death rate was approximately double in males than in females of the same age, and appears higher than the male mortality excess observed for annual mortality rates unrelated to COVID-19 (M/F mortality ratio from 1.6 to 1.7). Interestingly, this excess mortality in males was identical when considering suspected and confirmed cases of COVID-19 deaths in care homes (results not shown), suggesting that most suspected cases were correctly diagnosed. This is also supported by the very good quantitative correspondence between the daily deaths attributed to COVID-19 (including suspected cases) and the daily deaths excess occurring in spring 2020 compared to previous years (Molenberghs *et al*. 2020; Wu *et al*. 2020). Despite their higher vulnerability, males constituted only 45.1% of COVID-19 deaths in Wallonia, a paradox resulting from the much lower proportion of males (3.9%) than females (9.2%) ≥65 years old living in care homes, while the care homes population has been more infected.

We tentatively interpreted the higher ratio of COVID-19 over annual death rates in care homes compared to the UA population as reflecting a higher contamination in care homes, due to the potentially rapid spread of the virus in such an environment (Arons *et al*. 2020). However, other factors may also play a role, in particular the quality of care treatments, given that a minority of care home residents who died from COVID-19 were hospitalized (28%) whereas nearly all the victims in the UA population were hospitalized. Moreover, during the peak of the death wave, in particular between April to early May, the proportion of deaths of care home residents in hospital was lower (26%) than just before and after (41%), possibly because the risk of saturating hospitals’ intensive care facilities influenced the decision to hospitalize or not COVID-19 patients. Nevertheless, in hospitals, the proportion of patients of whatever origin who died from COVID-19 was very high among the ≥80 years old (39%) compared to the <60 years old (4%) (Van Beckhoven *et al*., 2020), so that even if a higher proportion of care home residents were hospitalized, it may not have changed much their survival chance. Moreover, differences in care quality cannot explain why HEP, in which 40% of the residents who died from COVID-19 were hospitalized, were 2.4 times less affected than NH institutions, even after factoring out the difference in health condition of their respective populations. The lower level of close contacts with caregivers in HEP compared to NH is a more parsimonious explanation of this contrast.

We currently lack serological tests data allowing to compare the level of presumed infection in the different populations but for the general population in Belgium, seroprevalence reached 6.0% (95% CI 5.1 to 7.1; Herzog *et al*. 2020) in the week of 20-26 April 2020, a value similar to that reported in other well-hit countries (Eckerle & Meyer 2020). If we assume that the contamination by the SARS-CoV-2 virus has been 3.5 more prevalent in care homes than outside them, we could expect a seroprevalence of c. 20% in care homes. This seems not incompatible with the systematic PCR tests campaign performed in nursing homes since April 8 which reported that 9% (21,979 tests until 28 April) and then 5% (50,736 tests until May 21) of the residents of care homes in Wallonia were positive (Sciensano 2020a & 2020b), given that c. 4% of the care home population had already died from COVID-19 when these tests were performed, and a significant percentage of the residents might have already fully recovered from viral infection when they were tested because the viral load of SARS-CoV-2 typically becomes undetectable about two weeks after symptom onset (Walsh *et al*. 2020). It is also worth noting that c.75% of the positively tested care home residents were asymptomatic or possibly pre-symptomatic (Hoxha *et al*. 2020), a figure reported in other studies (e.g. Ladhani et al. 2020).

Other lines of evidence of the importance of viral transmission in care homes come from (i) the temporal dynamics of deaths and (ii) the distribution of COVID-19 deaths and cases per institution. First, the delayed but then faster increase of deaths in care homes compared to the external population (Fig. 2) is consistent with primary infections originating from the care home personal or possibly visitors (explaining the delay), followed by a more rapid viral contamination within each institution contaminated due to the difficulty to limit interpersonal contacts and/or to the lack of PPE (explaining the faster increase).

Second, the crude COVID-19 death rate was very heterogeneous among care home institutions, with 54% of them having no COVID-19 death to deplore, while 17% of them lost at least 100‰ of their residents, and 2.6‰ lost between 200‰ and up to 340‰ of their residents. If we assume that in the latter most hit institutions virtually all the residents had been infected and led to an average mortality rate of 250‰, a mean mortality rate of 44.2‰ over all institutions would correspond to a prevalence of 44.2/250 ≈ 18%. Hence, a different reasoning suggest again that about a fifth of the residents of care homes had been contaminated by SARS-CoV-2. The absence of COVID-19 deaths mostly occurred in HEP (without care facilities) and in small institutions, while high mortality rates mostly occurred in NH and tended to increase with the bed capacity of the institution (Fig. 1). Hence, the risk that the SARS-SoV-2 virus entered and spread through a care home clearly increased with the presence of a health care staff and with the size of the institution, as expected from the potential number of interpersonal contacts, during a period where PPE and possibly adequate staff formation for such epidemic were in deficit. It is worth noting that the nearly two-fold lower mortality rate observed in medium-sized HEP (2.1%) compared to NH (4.0%) is very consistent with the distribution of positive COVID-19 tests performed from end April to May 2020 (Hoxha *et al*. 2020). Among institutions where at least 50% of the residents and staff were tested, the rate of positive tests reached 2.5% for residents and 1.6% for staff in HEP (7,402 persons tested in 100 institutions), against 5.9% for residents and 4.2% for staff in NH (60,667 persons tested in 370 institutions; unpublished results). The importance of transmission by staff was also highlighted in six cares homes of London, where SARS-CoV-2 genome sequencing showed that there were often multiple introductions of the virus per institution and that staff working across different care homes had a 3-fold higher risk of being contaminated than staff working in single care homes (Ladhani et al. 2020). Conversely, in France, 17 nursing homes where the staff decided to self-confine voluntarily with the residents at the beginning of the pandemic recorded 4 to 8-times less cases and deaths than the national averages (Belmin et al. 2020).

## Conclusion

Care homes represent a very specific context that has proven to be highly impacted by the COVID-19 epidemic, due both to the particular vulnerability of its population and the difficulty to contain the SARS-CoV-2 transmission once it has infected an institution. Given the high death toll that care homes underwent in many Western countries, care homes should be given special attention to understand how to avoid primary infection and how to limit contamination of its residents and staff. Our global analyses of death rates revealed that the size of the institution and the importance of nursing services provided (higher in NH than in HEP) were important factors determining the relative death toll. Nevertheless, our analyses do not reveal why nearly half of the institutions have not recorded any death while nearly one in six lost at least a tenth of their residents. These contrasts may result from (i) the stochastic nature of a primary infection (as suggested by the institution size effect), (ii) differences in mean health status of residents among institutions (e.g. HEP *versus* NH), or (iii) differences in the organisation and/or specific measures taken by each institution in response to the COVID-19 epidemic. Further research investigating the history of infections within representative institutions could clarify these questions. A comparison of serologic tests conducted inside and outside care homes would also be helpful to confirm our interpretation that the prevalence of SARS-CoV-2 was much higher within care homes than outside them. Finally, we recommend that epidemiological models integrate care home populations as a specific entity in their forecast.

## Data Availability

N/A

## Acknowledgments

We thank Patrick Lusyne from StatBel for helping us obtain the adequate population and annual deaths data, and for interesting discussions to interpret our results. We also warmly thank the staff of the hospitals and care homes for their conscientious reporting of COVID-19 deaths despite the difficult times. OJH, SD and TE are supported by the *Fonds de la Recherche Scientifique* (F.R.S. - FNRS, Belgium).

## Footnotes

1 The terms used here to designate different categories of care homes are those indicated on the Healthy Belgium website of the Belgian federal government (https://www.healthybelgium.be/en/health-system-performance-assessment/specific-domains/care-for-the-elderly)

## References

Armitage, R., & Nellums, L. B. (2020). COVID-19 and the consequences of isolating the elderly. The Lancet Public Health, 5(5), e256. https://doi.org/10.1016/S2468-2667(20)30061-X

Arons, M. M., Hatfield, K. M., Reddy, S. C., Kimball, A., James, A., Jacobs, J. R., Taylor, J., Spicer, K., Bardossy, A. C., Oakley, L. P., Tanwar, S., Dyal, J. W., Harney, J., Chisty, Z., Bell, J. M., Methner, M., Paul, P., Carlson, C. M., McLaughlin, H. P.,… Public Health–Seattle and King County and CDC COVID-19 Investigation Team. (2020). Presymptomatic SARS-CoV-2 Infections and Transmission in a Skilled Nursing Facility. The New England Journal of Medicine, 382(22), 2081-2090. https://doi.org/10.1056/NEJMoa2008457

Ars P., Dal L., Poulain M. (1988) Comment appréhender le problème statistique des petits nombres en démographie ?, in Les migrations internationales (Actes du colloque de l’AIDELF de Calabre, 1986), pp. 156-170

Belmin, J., Um-Din, N., Donadio, C., Magri, M., Nghiem, Q. D., Oquendo, B., Pariel, S., Lafuente-Lafuente, C. (2020). Coronavirus Disease 2019 Outcomes in French Nursing Homes That Implemented Staff Confinement With Residents. JAMA Network Open, 3(8), e2017533-e2017533. https://doi.org/10.1001/jamanetworkopen.2020.17533

Bustos Sierra N, Bossuyt N, Braeye T, et al. All-cause mortality supports the COVID-19 mortality figures in Belgium. Submitted for publication.

Comas-Herrera A, Ashcroft E and Lorenz-Dant K. (2020) International examples of measures to prevent and manage COVID-19 outbreaks in residential care and nursing home settings. Report in LTCcovid.org, International Long-Term Care Policy Network, CPEC-LSE, 11 May 2020. https://ltccovid.org/wp-content/uploads/2020/05/International-measures-to-prevent-and-manage-COVID19-infections-in-care-homes-2-May-1.pdf

Comas-Herrera A, Zalakaín J, Litwin C, Hsu AT, Lemmon E, Henderson D and Fernández J-L (2020) Mortality associated with COVID-19 outbreaks in care homes: early international evidence. Article in LTCcovid.org, International Long-Term Care Policy Network, CPEC-LSE, 26 June 2020. https://ltccovid.org/wp-content/uploads/2020/06/Mortality-associated-with-COVID-among-people-who-use-long-term-care-26-June-1.pdf

Dequeker, S., Latour, K., Vandaele E., Islamaj, E., Int Panis, L. (2020). COVID-19 surveillance in residential institutions. Version 3.3-01/07/2020. https://www.sciensano.be/sites/default/files/protocol_covid-19_surveillance_in_residential_institutions_20200701_version_3.3.pdf

Dolejs, J. (2015). Age Trajectories of Mortality from All Diseases in Five Countries of Central Europe During the Last Decades. Biodemography and Social Biology, 61(1), 40-64. https://doi.org/10.1080/19485565.2014.936999

Eckerle, I., & Meyer, B. (2020). SARS-CoV-2 seroprevalence in COVID-19 hotspots. The Lancet. https://doi.org/10.1016/S0140-6736(20)31482-3

ECDC Public Health Emergency Team, Danis Kostas, Fonteneau Laure, Georges Scarlett, Daniau Côme, Bernard-Stoecklin Sibylle, Domegan Lisa, O’Donnell Joan, Hauge Siri Helene, Dequeker Sara, Vandael Eline, Van der Heyden Johan, Renard Françoise, Sierra Natalia Bustos, Ricchizzi Enrico, Schweickert Birgitta, Schmidt Nicole, Abu Sin Muna, Eckmanns Tim, Paiva José-Artur, Schneider Elke. High impact of COVID-19 in long-term care facilities, suggestion for monitoring in the EU/EEA, May 2020. Euro Surveill. 2020;25(22):pii=2000956. https://doi.org/10.2807/1560-7917.ES.2020.25.22.2000956

Einiö, E., Guilbault, C., Martikainen, P. & Poulain, M. (2012). Gender Differences in Care Home Use among Older Finns and Belgians. Population, vol. 67(1), 71-95. https://doi.org:10.3917/popu.1201.0075

Falconer, M., & O’Neill, D. (2007). Profiling disability within nursing homes: a census-based approach. Age Ageing, 36(2), 209-213. https://doi.org/10.1093/ageing/afl185

Fisman, D. N., Bogoch, I., Lapointe-Shaw, L., McCready, J., & Tuite, A. R. (2020). Risk Factors Associated With Mortality Among Residents With Coronavirus Disease 2019 (COVID-19) in Long-term Care Facilities in Ontario, Canada. JAMA Network Open, 3(7), e2015957-e2015957. https://doi.org/10.1001/jamanetworkopen.2020.15957

Flaxman, S., Mishra, S., Gandy, A., Unwin, H. J. T., Mellan, T. A., Coupland, H., Whittaker, C., Zhu, H., Berah, T., Eaton, J. W., Monod, M., Ghani, A. C., Donnelly, C. A., Riley, S., Vollmer, M. A. C., Ferguson, N. M., Okell, L. C., & Bhatt, S. (2020). Estimating the effects of non-pharmaceutical interventions on COVID-19 in Europe. Nature, 584(7820), 257-261. https://doi.org/10.1038/s41586-020-2405-7

Gompertz, B. (1825). XXIV. On the nature of the function expressive of the law of human mortality, and on a new mode of determining the value of life contingencies. In a letter to Francis Baily, Esq. F. R. S. &c. Philosophical Transactions of the Royal Society of London, 115, 513-583. https://doi.org/10.1098/rstl.1825.0026

Herm, A., Poulain, M., & Anson, J. (2014). Excess mortality risks in institutions: The influence of health and disability status. In Mortality in an International Perspective (pp. 245-263). Springer, Cham.

Herzog, S., Bie, J. D., Abrams, S., Wouters, I., Ekinci, E., Patteet, L., Coppens, A., Spiegeleer, S. D., Beutels, P., Damme, P. V., Hens, N., & Theeten, H. (2020). Seroprevalence of IgG antibodies against SARS coronavirus 2 in Belgium: A prospective cross-sectional study of residual samples. *MedRxiv*, https://doi.org/10.1101/2020.06.08.20125179

Horiuchi, S., Finch, C. E., Meslé, F., & Vallin, J. (2003). Differential patterns of age-related mortality increase in middle age and old age. The Journals of Gerontology Series A: Biological Sciences and Medical Sciences, 58(6), B495-B507. https://doi.org/10.1093/gerona/58.6.B495

Hoxha, A., Wyndham-Thomas, C., Klamer, S., Dubourg, D., Vermeulen, M., Hammami, N., & Cornelissen, L. (2020). Asymptomatic SARS-CoV-2 infection in Belgian long-term care facilities. The Lancet Infectious Diseases. https://doi.org/10.1016/S1473-3099(20)30560-0

Humblet, P. (2020). Inégalités sociales de santé: Tout a changé ? Politique, 112, 99-102

Imaizumi, Y. (1996). Longitudinal analysis of mortality from breast cancer in Japan, 1950-1993: fitting Gompertz and Weibull functions. Mechanisms of ageing and development, 88(3), 169-183. https://doi.org/10.1016/0047-6374(96)01735-6

Ladhani, S. N., Chow, J. Y., Janarthanan, R., Fok, J., Crawley-Boevey, E., Vusirikala, A., Fernandez, E., Perez, M. S., Tang, S., Dun-Campbell, K., Evans, E. W.-, Bell, A., Patel, B., Amin-Chowdhury, Z., Aiano, F., Paranthaman, K., Ma, T., Saavedra-Campos, M., Myers, R.,… Ramsay, M. E. (2020). Increased risk of SARS-CoV-2 infection in staff working across different care homes enchanced CoVID-19 outbreak investigations in London care Homes. The Journal of Infection (in press). https://doi.org/10.1016/Minf.2020.07.027

Laferrère, A., Van den Heede, A., Van den Bosch, K., & Geerts, J. (2013). 22 Entry into institutional care: predictors and alternatives. In Active ageing and solidarity between generations in Europe: First results from SHARE after the economic crisis, (pp. 253-264). DeGruyter.

Liu, T., Wu, S., Tao, H., Zeng, G., Zhou, F., Guo, F., & Wang, X. (2020). Prevalence of IgG antibodies to SARS-CoV-2 in Wuhan—Implications for the ability to produce long-lasting protective antibodies against SARS-CoV-2. *MedRxiv*, 2020.06.13.20130252. https://doi.org/10.1101/2020.06.13.20130252

Logar, S. (2020). Care home facilities as new COVID-19 hotspots: Lombardy Region (Italy) case study. Arch Gerontol Geriatr. 89:104087. https://doi:10.1016/j.archger.2020.104087

Molenberghs, G., Faes, C., Aerts, J., Theeten, H., Devleesschauwer, B., Bustos Sierra, N., Braeye, T., Renard, F., Herzog, S., Lusyne, P., Van der Heyden, J., Van Oyen, H., Van Damme, P., & Hens, N. (2020). Belgian COVID-19 Mortality, Excess Deaths, Number of Deaths per Million, and Infection Fatality Rates (8 March—9 May 2020) [Preprint]. Epidemiology. https://doi.org/10.1101/2020.06.20.20136234

Petretto, D.R.; Pili, R. (2020). Ageing and COVID-19: What Is the Role for Elderly People? Geriatrics, 5, 25.

Quigley, D. D., Dick, A., Agarwal, M., Jones, K. M., Mody, L., & Stone, P. W. (2020). COVID-19 Preparedness in Nursing Homes in the Midst of the Pandemic. Journal of the American Geriatrics Society. 2020 May 12: 10.1111/jgs.16520. https://doi:10.1111/jgs.16520

Rada, A. G. (2020). Covid-19: the precarious position of Spain’s nursing homes. BMJ, 369:m1586. doi:https://doi.org/10.1136/bmj.m1586

Ricklefs, R. E., & Scheuerlein, A. (2002). Biological implications of the Weibull and Gompertz models of aging. Journals of Gerontology - Series A Biological Sciences and Medical Sciences, 57(2), B69-B76. Scopus. https://doi.org/10.1093/gerona/57.2.B69

Riggs, J. E. (1991b). Longitudinal Gompertzian analysis of ischemic heart disease mortality in the US, 1962-1986: a method of demonstrating the deterministic dynamics describing its decline. Mechanisms of ageing and development, 57(1), 1-14. https://doi.org/10.1016/0047-6374(91)90020-Z

Robbiani, D. F., Gaebler, C., Muecksch, F., Lorenzi, J. C. C., Wang, Z., Cho, A., Agudelo, M., Barnes, C. O., Gazumyan, A., Finkin, S., Hägglöf, T., Oliveira, T. Y., Viant, C., Hurley, A., Hoffmann, H.-H., Millard, K. G., Kost, R. G., Cipolla, M., Gordon, K.,… Nussenzweig, M. C. (2020). Convergent antibody responses to SARS-CoV-2 in convalescent individuals. Nature, 1-8. https://doi.org/10.1038/s41586-020-2456-9

Sciensano (2020a). COVID-19 Bulletin Epidémiologique Hebdomadaire du 30 avril 2020, Bruxelles, Belgique. https://covid-19.sciensano.be/sites/default/files/Covid19/COVID-19_Weekly%20report_20200430%20-%20FR_0.pdf

Sciensano (2020b). COVID-19 Bulletin Epidémiologique Hebdomadaire du 22 mai 2020, Bruxelles, Belgique. https://covid-19.sciensano.be/sites/default/files/Covid19/COVID-19_Weekly%20report_20200522%20-%20FR.pdf

Sciensano (2020c). COVID-19 Bulletin Epidémiologique Hebdomadaire du 19 juin 2020, Bruxelles, Belgique. https://covid-19.sciensano.be/sites/default/files/Covid19/COVID-19Weekly%20report20200619%20-%20FR.pdf

Szczerbińska, K. (2020). Could we have done better with COVID-19 in nursing homes? Eur Geriatr Med. https://doi.org/10.1007/s41999-020-00362-7

Trabucchi, M., & De Leo, D. (2020). Nursing homes or besieged castles: COVID-19 in northern Italy. The Lancet Psychiatry, 7(5), 387-388. https://doi.org/10.1016/S2215-0366(20)30149-8

Van Beckhoven D., Duysburgh E., Montourcy M., De Rouck M., Vilain A. Catteau L. Deblonde J., Wyndham-Thomas C., Van Goethem N. (2020). Points clés de la surveillance des patients hospitalisés atteints d’une infection COVID-19 confirmée: Résultats jusqu’au 14 juin 2020. Bruxelles, Belgique: Sciensano. Numéro de dépôt légal: D/2020/14.440/65. https://covid-19.sciensano.be/sites/default/files/Covid19/COVID-19THEMATIC%20REPORTCOVID-19%20HOSPITALISED%20PATIENTSFR.pdf

Verbeek, H., Gerritsen, D. L., Backhaus, R., de Boer, B. S., Koopmans, R. T., & Hamers, J. P. (2020). Allowing visitors back in the nursing home during the COVID-19 crisis–A Dutch national study into first experiences and impact on well-being. Journal of the American Medical Directors Association. 21(7), 900-904. https://doi.org/10.1016/Mamda.2020.06.020

Verity, R., Okell, L. C., Dorigatti, I., Winskill, P., Whittaker, C., Imai, N., Cuomo-Dannenburg, G., Thompson, H., Walker, P. G. T., Fu, H., Dighe, A., Griffin, J. T., Baguelin, M., Bhatia, S., Boonyasiri, A., Cori, A., Cucunubá, Z., FitzJohn, R., Gaythorpe, K.,… Ferguson, N. M. (2020). Estimates of the severity of coronavirus disease 2019: A model-based analysis. The Lancet Infectious Diseases, 20(6), 669-677. https://doi.org/10.1016/S1473-3099(20)30243-7

Walsh, K. A., Jordan, K., Clyne, B., Rohde, D., Drummond, L., Byrne, P., Ahern, S., Carty, P. G., O’Brien, K. K., O’Murchu, E., O’Neill, M., Smith, S. M., Ryan, M., & Harrington, P. (2020). SARS-CoV-2 detection, viral load and infectivity over the course of an infection. The Journal of Infection. https://doi.org/10.1016/Minf.2020.06.067

Wenham, C., Smith, J., & Morgan, R. (2020). COVID-19: The gendered impacts of the outbreak. The Lancet, 395(10227), 846-848. https://doi.org/10.1016/S0140-6736(20)30526-2

Williamson, E. J., Walker, A. J., Bhaskaran, K., Bacon, S., Bates, C., Morton, C. E., Curtis, H. J., Mehrkar, A., Evans, D., Inglesby, P., Cockburn, J., McDonald, H. I., MacKenna, B., Tomlinson, L., Douglas, I. J., Rentsch, C. T., Mathur, R., Wong, A. Y. S., Grieve, R.,… Goldacre, B. (2020). OpenSAFELY: Factors associated with COVID-19 death in 17 million patients. Nature, 1-11. https://doi.org/10.1038/s41586-020-2521-4

Wu, J., McCann, A., Katz, J., & Peltier, E. (2020). 153,000 Missing Deaths: Tracking the True Toll of the Coronavirus Outbreak. The New York Times. Retrieved 22 July 2020, from https://www.nytimes.com/interactive/2020/04/21/world/coronavirus-missing-deaths.html

